# Tracking and forecasting milepost moments of the epidemic in the early-outbreak: framework and applications to the COVID-19

**DOI:** 10.1101/2020.03.21.20040139

**Authors:** Huiwen Wang, Yanwen Zhang, Shan Lu, Shanshan Wang

**Author notes:** Corresponding author. Correspondence to: School of Economics and Management, Beihang University, Beijing 100191, China. E-mail address (S.S. Wang).

## Abstract

**Background:** The outbreak of the 2019 novel coronavirus (COVID-19) has attracted global attention. In the early stage of the outbreak, the most important question concerns some meaningful milepost moments, including (1) the time when the number of daily confirmed cases decreases, (2) the time when the number of daily confirmed cases becomes smaller than that of the daily removed (recovered and death), (3) the time when the number of daily confirmed cases becomes zero, and (4) the time when the number of patients treated in hospital is zero, which indicates the end of the epidemic. Intuitively, the former two can be regarded as two important turning points which indicate the alleviation of epidemic to some extent, while the latter two as two “zero” points, respectively. Unfortunately, it is extremely difficult to make right and precise prediction due to the limited amount of available data at a early stage of the outbreak.

**Method:** To address it, in this paper, we propose a flexible framework incorporating the effectiveness of the government control to forecast the whole process of a new unknown infectious disease in its early-outbreak. Specially, we first establish the iconic indicators to characterize the extent of epidemic spread, yielding four periods of the whole process corresponding to the four meaningful milepost moments: two turning points and two “zero” points. Then we develop the tracking and forecasting procedure with mild and reasonable assumption. Finally we apply it to analyze and evaluate the COVID-19 using the public available data for mainland China beyond Hubei Province from the China Centers for Disease Control (CDC) during the period of Jan 29th, 2020, to Feb 29th, 2020, which shows the effectiveness of the proposed procedure.

**Results:** Results show that our model can clearly outline the development of the epidemic at a very early stage. The first prediction results on Jan 29th reveal that the first and second milepost moments for mainland China beyond Hubei Province would appear on Jan 31st and Feb 14th respectively, which are only one day and three days behind the real world situations. Forecasting results indicate that the number of newly confirmed cases will become zero in the mid-late March, and the number of patients treated in the hospital will become zero between mid-March and mid-April in mainland China beyond Hubei Province. The framework proposed in this paper can help people get a general understanding of the epidemic trends in counties where COVID-19 are raging as well as any other outbreaks of new and unknown infectious diseases in the future.

## 1 Introduction

The atypical pneumonia case caused by 2019 novel coronavirus (COVID-19), which is a highly infectious human disease, was first reported in Dec 31st, 2019 in Wuhan, the capital of Hubei Province in China (Organization et al., 2020). To mitigate the effect of epidemics spreading across China and other countries, Wuhan was temporarily shut-down from Jan 23th, 2020, which has proved to be efficient in timely stopping the spread of the coronavirus (Chinazzi et al., 2020). However, due to the “Spring Festival travel rush”, there was still a rising number of confirmed cases in China in the following two months, which has caused great challenges to medical resources (Li et al., 2020).

The questions that draw the most concerns are how COVID-19 will spread, and when it will end. People were always asking when the number of the daily confirmed cases will become smaller than the previous days, and when the daily confirmed cases will become smaller than that of the removed (recovered and death). These are not only of highly important for the general public, but also for government, who plays an important role in controlling the disease within a short period as much as possible. Since the decline of the number of newly confirmed cases and the number of patients in hospital imply the alleviation of epidemic, the emergences of their turning points convey useful information for decision making on medical resources allocation and isolation policies in the post-stage of the epidemic.

Meanwhile, it is also important to predict when will the number of daily confirmed cases become zero, as well as when the number of infectious cases in hospital will be zero. The latter indicates the end of the epidemic. These two “zero points” can also help the government to consider loosing population migration restriction among cities. Additionally, authorities in economic departments can use the forecasting results to assess the impact of the epidemic on the economy in advance, and making planning for the restoration of normal production and living order.

There have been various literatures on COVID-19 from different aspects, i.e., the origin of COVID-19, the clinical features as well as epidemic transmission characteristics. Specifically, for the origin of the virus, (Fan et al., 2019) and (Luk et al., 2019) pointed out that COVID-19 is an infectious disease caused by a virus closely related to SARS-CoV-2, while others believed that the COVID-19 virus was originally derived from wild animals (Huang et al., 2020; Benvenuto et al., 2020). For the epidemic transmission characteristics, Holshue et al. (2020) and Hui et al. (2020) found that the virus can be transmitted from person to person and that it has a high interpersonal transmission rate. Zhao et al. (2020) investigated the preliminary estimation of the basic reproduction number *R*_0_, which ranges 2.24(95%CI: 1.96 − 2.55) to 3.58(95%CI: 2.89 − 4.39) in the early outbreak, while Prasse et al. (2020) estimated it around 2.2, Tang et al. (2020) applied likelihood-based and model-based methods to the analysis of early reported cases, and the results showed that *R*_0_ is as high as 6.47. Zhou et al. (2020) used the SEIR model and stated that the range of *R*_0_ of COVID-19 is 2.8-3.3, indicating that the early pathogenic transmission capacity of COVID-19 is close to or slightly higher than SARS. Other literatures related *R*_0_ are Anastassopoulou et al. (2020); Zhang et al. (2020) and reference therein. Unfortunately, each of these models may results in different estimations of *R*_0_, which may causes any predictions based on *R*_0_ unstable.

Recently, lots of literatures are related to the trend prediction of the COVID-19 in China. Zeng et al. (2020) proposed a multi-model ordinary differential equation set neural network and model-free methods to predict the interprovincial transmissions in mainland China, especially those from Hubei Province, and predicted that the COVID-19 in China is likely to decelerate before Feb 18th and to end before April 2020. Chen et al. (2020) made prediction based on epidemiological surveys and analyses, which showed that the total number of diagnoses would be 2-3 times that of SARS, and the peak is predicted to be in early or middle February. Yu et al. (2020) revised the SIR model based on the characteristics of the COVID-19 epidemic development, and proposed a time-varying parameter-SIR model to study the trend of the number of infected people. Peng et al. (2020) used the SEIR method to predict the end of the epidemic in most cities in mainland China. Wu et al. (2020) used the Markov chain Monte Carlo method to estimate *R*_0_, and inferred from the SEIR model that the peak COVID in Wuhan would be reached in April, and other cities in China would be delayed by 1 to 2 weeks.

However, there are some obvious shortcomings of forecasting method based on epidemic model in terms of outbreak prediction. For example, SEIR model is a mathematical method relying on an assumption of epidemiological parameters for disease progression, which is absent for the novel pathogen. For instance, the basic infection number *R*_0_, the daily recovery rate, the characteristics of the disease itself (such as the infection rate and the conversion rate of the latent to the infected), the daily exposure rate of the latent and infected, and their initial population infection status (total population, infected, the initial value of the latent, the susceptible, the healer, etc.) and many other key parameters need to be set. For infectious diseases that have already appeared in the past, or those who have a large amount of data, it is not difficult to obtain these parameters. However, for unknown, sudden and early infectious diseases, obtaining these parameters is full of difficulties, which leads to a great uncertainty and limitations in the prediction of the epidemic situation using the SEIR model.

Moreover, there exist many challenges for the prediction of a new epidemic situation similar to the COVID-19. First, little prior knowledge can be used to analogize or refer to for a brand new epidemic; secondly, the existence of government management will make the development of the epidemic completely different from that under free development, thus how to incorporate the influence of government measures into the fitting process of parameters and build a statistical model from this should be taken into consideration; thirdly, in the early-outbreak the initial data often fluctuates violently and the data quality is low, thus many commonly used parameter estimation methods are not applicable anymore; furthermore, the amount of data in early stage is too small, so it is difficult to directly rely on the inertia of the data to make forward prediction. In summary, in the early stages of brand new epidemics, how to use some low-quality and small data sets to make basic and relatively accurate forecast judgments for the entire process of the epidemic, is a long-term pain point.

To cope with these challenges, we propose a simple and effective framework incorporating the effectiveness of the government control to forecast the whole process of a new unknown infectious disease in its early-outbreak, from which we emphasis on the prediction of meaningful milepost moments. Specifically, we first propose a series of iconic indicators to characterize the extent of epidemic spread, and describe four periods of the whole process corresponding to the four meaningful milepost moments: two turning points and two “zero” points; then we develop the proposed procedure with mild and reasonable assumption, especially without relying on an assumption of epidemiological parameters for disease progression. Finally we apply it to analyze and evaluate the COVID-19 using the public available data in mainland China beyond Hubei Province from the China CDC during the period of Jan 29th, 2020, to Feb 29th, 2020, which shows the effectiveness of the proposed procedure. From the empirical study, we can conjecture that the proposed method may cast a flexible framework and perspective for early prediction of a sudden and unknown new infectious disease with effective government control. Specifically, in the early stage of the epidemic when some regular information is initially displayed, the proposed method can be used to predict the process of epidemic development and to judge which stage of development the situation is at, when the peak will be reached, and when the turning point will appear. Moreover, by continuously accumulating data and updating the model during the development of the epidemic, we can also predict when the epidemic will basically end. Finally, the proposed method enjoys great generalizability, which can be generalized to understand the epidemiological trend of COVID-19 spread in other counties, which will provide useful guidance for fighting against it.

The reminder of this paper is organized as follows. In Section 2, we proposed the main methodology, where we defined the iconic indicators to characterize the extent of epidemic spread in Section 2.1, yielding four periods of the whole process corresponding to the four meaningful milepost moments: two turning points and two “zero” points in Section 2.2, then Section 2.3 presents the proposed procedure with mild and reasonable assumption. Then we applied the proposed method to the COVID-19 using the public available data in mainland China beyond Hubei Province from the China CDC during the period of Jan 29th, 2020, to Feb 29th, 2020, and describe the trend of the COVID-19 spread in detail in Section 3. Some conclusions and discussions are finally given in Section 4.

## 2 Methodology

In order to assess and predict the epidemic, we first define a set of necessary indicators that can reflect the status of disease contagion. We then divide the cycle of epidemic into four stages, which divided by the turning points of the proposed indicators. Finally, we propose a computational framework to predict the turning points.

### 2.1 The iconic indicators to characterize a epidemic

It is obvious that the contagion process of an unknown virus in different regions would be diverse with respect to the number of patients and the growth pattern of epidemic, because of population density, population mobility, public health conditions, disease prevention and control measures. Therefore, we first constructed a set of indicators to monitor the essential laws of the development of the disease.

There are several requirements for the monitoring indicators. Firstly, the scale of the data should be eliminated so that the analysis methods and results are comparable across regions. Secondly, they can well reflect the general laws and characteristics of the epidemic process as well as accurately and coherently describe the entire process of the epidemic from the begin to the end. Especially, they should be able to answer the question of when the turning point of the epidemic would appear. Thirdly, they should be as simple and convenient as possible so that it can be applied with publicly available data. Last but not the least, the indicators should have clear meaning and good interpretability.

Following the above, we first adopt three basic indicators that are published daily by the provincial and municipal governments of China. That is, for time *t*, the daily confirmed cases *E*_*t*_, the daily recovered ones *O*_*t*_, the daily deaths *D*_*t*_. Then we define a few monitoring indicators to characterize the epidemic stages, that is the number of infectious cases in hospital *N*_*t*_, the daily infection rate *K*_*t*_ and the daily removed (the sum of recovered and deaths) rate *I*_*t*_, which are defined as follows.

- The number of infectious cases in hospital *N*_*t*_ is defined as the cumulative confirmed cases with recovered ones and deaths removed up to *t*, that is

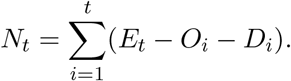 Note that *N*_*t*_ is essential for epidemic investigation, since it reflects the size of local patients and the pressure of medical system.
- The daily infection rate *K*_*t*_ is defined as the ratio of the daily confirmed cases at time *t* and the number of infectious cases in hospital at time *t* − 1, i.e.

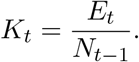 Obviously, *K*_*t*_ reflects the rate at which patients enter the treatment system. It is influenced by many factors, including the property of infectious diseases, the average immune capacity of the population, population density, climate condition, public health conditions, public health awareness, the awareness of self-prevention of diseases and the efforts of epidemic prevention and control.
- Similarly, the daily removed rate *I*_*t*_ is defined as the ratio of the daily removed cases at time *t* and the number of infectious cases in hospital at time *t* − 1, i.e.

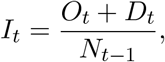

where *I*_*t*_ reflects the rate at which patients leave the medical system, that is, the rate at which the pressure of medical resource is released. Using the above indicators, we further define *R*_*t*_ as the outbreak status on day *t* as follow:

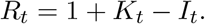

Obviously, it holds that

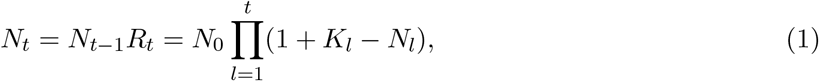

where *N*_0_ denotes the initial number of patients in hospital at the beginning of the outbreak. In particular, when the daily infection rate and removed rate are relatively stable, denoted as *K* and *I* respectively, we have the constant epidemic status index *R* = 1 + *K* − *I*. Then (1) can be written as:

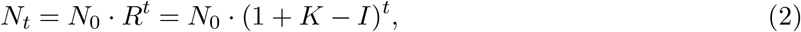

which shows that the epidemic situation is in the form of an exponential curve. And the epidemic status indicator *R* can well reflect the rate of expansion or convergence of the population with infectious capacity.

### 2.2 Four stages of a epidemic

In this section, we will describe the whole process of a epidemic under assumption that the government has implemented effective control measures, which can be divided into four stages, i.e. “outbreak period”, “controlled period”, “mitigation period” and “convergence period” successively. And we will quantify the iconic features for each stage, which corresponds to the two turning points and two “zero” points, respectively.

#### Stage 1: Outbreak Period

In the initial stage of an epidemic outbreak, there is delay of social response due to the limited knowledge of the epidemic, and the power of contagion preventing and control is inevitably not enough. Thus the daily infection rate *K*_*t*_ would be high. At the same time, the healing process in the initial stage is relatively long, and the number of severe patients is small, leading the daily removed rate *I*_*t*_ to be close to zero. Therefore, the outbreak status indicator *R*_*t*_ during this period is usually much larger than 1, that is:

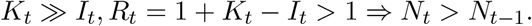

It can be seen that, during the outbreak period, the number of newly diagnosed patients increases sharply, and the number of patients in the hospital will increase dramatically correspondingly, which will pose great burden on medical institutions, especially for hospitals.

As the epidemic exacerbates, if the government began to intervene by a series of emergency measures, where a disease prevention and control system will be quickly established, the daily infection rate *K*_*t*_ will significantly decrease. Usually, the new daily confirmed cases will begin to decline as well. During the epidemic prevention and control process, once the situation improves, we will see the emergence of the first turning point denoted as *T*_1_. Then after the data *T*_1_, the newly diagnosed patients *E*_*t*_ changes from a rapid rise in the outbreak period to a descending channel (*E*_*t*_ < *E*_*t*−1_). In summary, the emergence of the first turning point *T*_1_ indicates that the disease control measures have begun to work, which implies the end of the “Outbreak Period”.

#### Stage 2: Controlled Period

The emergence of the first turning point is a very positive signal, indicating that the public health management measures have obviously taken effect and the epidemic has entered the “controlled period”. However, due to the fact that the completion rate *I*_*t*_ at this stage is still relatively low, the number of patients treated in the hospital will continue to increase. The controlled period will continue until the second turning point *T*_2_ appears, that is, patients in hospital *N*_*t*_ reaches the peak and starts to decline. This is because the completion rate increase so significantly that *K*_*t*_ = *I*_*t*_ is fulfilled after a long period of treatment in the previous stage. When the completion rate *I*_*t*_ surpasses infection rate *K*_*t*_, the number of patients treated in the hospital began to decline from peak.

#### Stage 3: Mitigation Period

The sign of the end of the controlled period is *K*_*t*_ = *I*_*t*_. Thereafter, *K*_*t*_ will continue to fall with the rise of *I*_*t*_, which gives

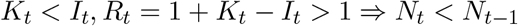

This indicates that the daily completion rate *I*_*t*_ will start to be greater than the daily infection rate *K*_*t*_, that is, the value of the outbreak status indicator *R*_*t*_ becomes less than 1. The population size with infectious capacity will be reduced, and the pressure of medical resources will be significantly relieved, marking the beginning of “mitigation period”. The mitigation period will continue until the appearance of zero report for newly confirmed cases, that is, *E*_*t*_ = 0, which we call it as the first “zero” point *Z*_1_. After the first zero point is reached, the intensity of prevention and control in the entire society will be relieved except for the hospital, that is, the “mitigation period” ends and the “convergence period” starts.

#### Stage 4: Convergence Period

The “convergence period” will end at the second “zero” point *Z*_2_, which means that the number of people treated in the hospital is equal to or close to zero. After reaching the second zero point, the epidemic is completely over.

For clarity, we summarize the iconic features and the corresponding milepost moments of each stage in the whole process of the epidemic in Table 1.

**Table 1:** The four stage of an epidemic

| Stage | Outbreak | Controlled | Mitigation | Convergence |
| --- | --- | --- | --- | --- |
| Begin<br>with | the number of newly<br>diagnosed increases | the number of newly<br>diagnosed decreases<br>(first turning point) | the number of<br>patients in hospital<br>decreases (second<br>turning point) | the number of newly<br>diagnosed equals to<br>0 (first zero point) |
| End<br>with | the number of newly<br>diagnosed reached<br>peak (first turning<br>point) | the number of<br>patients in hospital<br>reached peak<br>(second turning<br>point) | the number of<br>patients in hospital<br>equals to 0 (first<br>zero point) | the number of<br>patients in hospital<br>equals to 0 (second<br>zero point) |
| | $K \gg I, R \gg 1$ | $K > I, R > 1$ | $K < I, R < 1$ | $K = 0, R \ll 1$ |

### 2.3 Implementation: the proposed model

According to Section 2.2, the modeling and predicting of the epidemic need to be divided into two parts. The first part corresponds to the outbreak period, where the intervention and disease curing is not effective enough. The infection rate *K*_*t*_ increases rapidly and the completion rate *I*_*t*_ is small. Thus, the number of newly diagnosed patients *E*_*t*_ increases rapidly, and the number of patients treated in hospital *N*_*t*_ increases. The pressure on medical resources will soon be overwhelmed. According to equation (2), *N*_*t*_ will be in an exponential growth trend without forming a convex curve, nor will the so-called two turning points or two “zero” points appear.

The second part, which is the focus of this article, is when the *K*_*t*_ starts to decrease and *I*_*t*_ starts to increase due to effective intervention and improved healing level for individual patients. Only in this situation will the turning points and zero points *T*_1_, *T*_2_, *Z*_1_, *Z*_2_ successively appear, and then the epidemic could end. Therefore, we will model the development of the epidemic under the assumption of effective intervention, then we can obtain the early prediction of two turning points and two “zero” points based on the predicting modeling of *E*_*t*_ and *N*_*t*_.

Suppose that the infection rate *K*_*t*_ and the removed rate *I*_*t*_ change gently within a time window *m* before time *t*_0_ with exponential growth, then given *m* and *t*_0_, denote 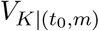 and 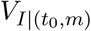 as the average change rate of *K*_*t*_ and *I*_*t*_ respectively, that is,

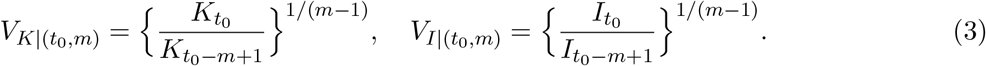

For any *t* > *t*_0_, the infection rate *K*_*t*_ and the removed rate *I*_*t*_ can be predicted as follows:

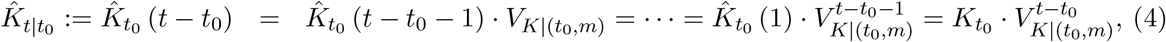

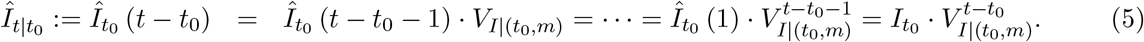

Thus, we can obtain the outbreak status *R*_*t*_, the number of patients in the hospital *N*_*t*_, and the number of newly diagnosed *E*_*t*_ as

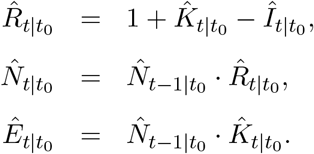

According to the prediction process, it can be seen that the prediction results mainly depend on 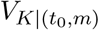 and 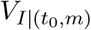, whose value is up to the selection of time window *m* and starting point *t*_0_. However, it is worth noticing that the selection of *m* and *t*_0_ is not arbitrary, which is suggested as in the follow assumption.

#### Assumption 1.

*The time window m and the starting point t*_0_ *should be chosen satisfying* 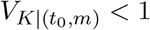 *and* 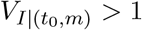. *Meanwhile, keeping I*_*t*_ < 1 *due to interpretability constraints, and the starting point t*_0_ *should be close to the date of the latest published data as much as possible*.

In summary, here we describe details of the proposed procedure in Algorithm 1.

#### Algorithm 1 Main Prediction Procedure

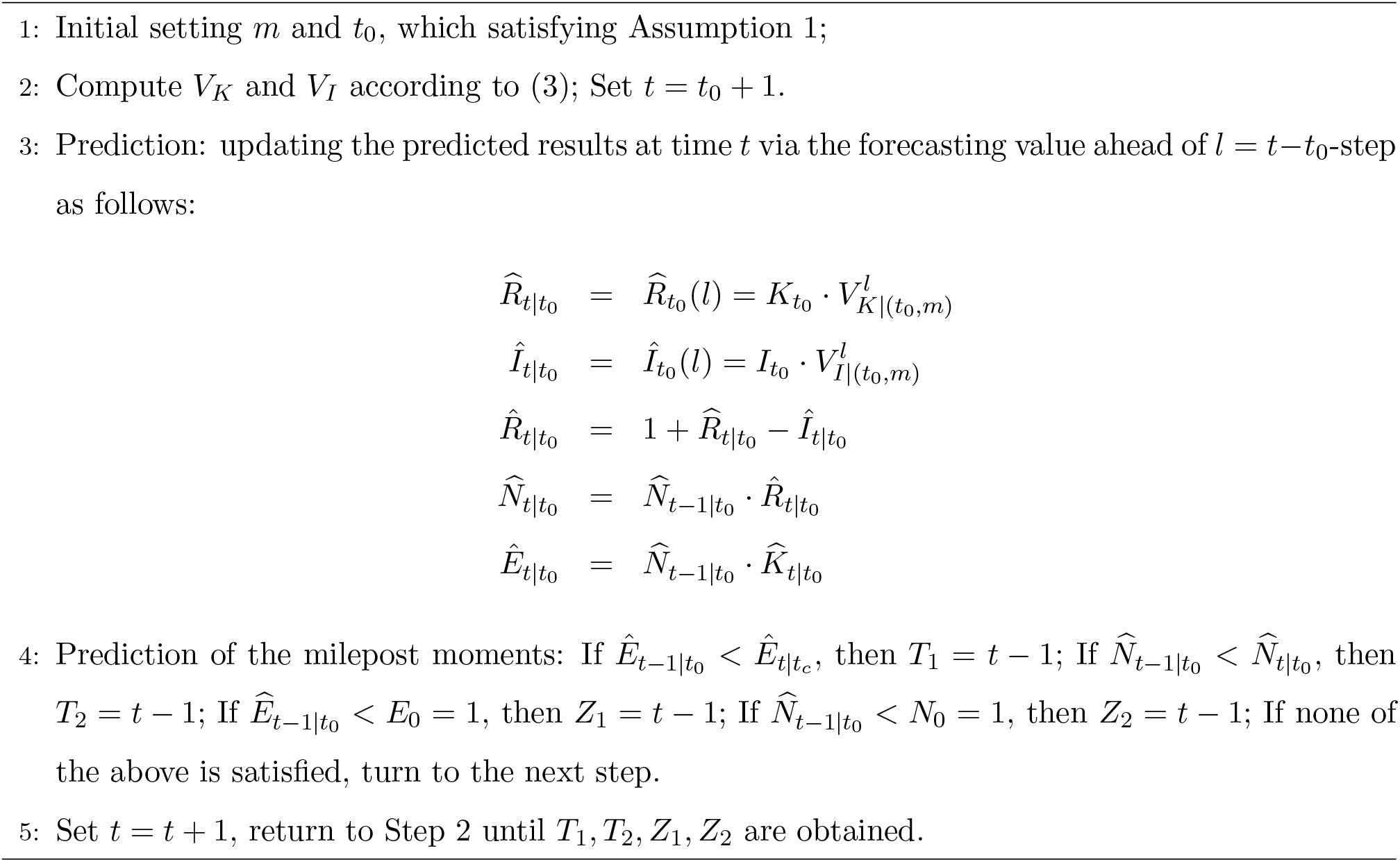

It is also worth noticing that in practice, the more data information we accumulate, the clearer the underly law of the epidemic. Therefore, we can also continuously modify the iterative prediction model according to the actual data, so that the prediction of the next stage and the prediction of the long-term situation can be more accurate.

## 3 Application: Analysis of the COVID-19 in mainland China beyond Hubei Province

We apply it to analyze and evaluate the COVID-19 using the public available data in mainland China beyond Hubei Province from the China CDC during the period of Jan 29th, 2020, to Feb 29th, 2020. Here we first show the actual trend of the COVID-19, and then compared with the predicted ones via the proposed method. Finally, we will show the effect of *m* on the predicted results.

### 3.1 The turning points and zero points observed

After the shutdown of most parts of Hubei province in Jan 23rd, other parts of China also immediately launched prevention and control strategies, including regional isolation, admission of all confirmed patients, isolating all suspected patients and so on. The effective implementation of these intervention policies quickly controlled the rapid spread of the epidemic in these areas. As can be seen in Figure 1, the parameter infectious rate *K*_*t*_, which reflects the intensity of the spread of the epidemic, has shown a significant downward trend since Jan 27th after severe fluctuations from Jan 22nd to 26th. As can be seen in Fig. 1, we find out that the daily confirmed cases reached at peak on Jan 30th, 2020, with 761 confirmed cases and then continued to decline for two consecutive days.

**Fig 1:**
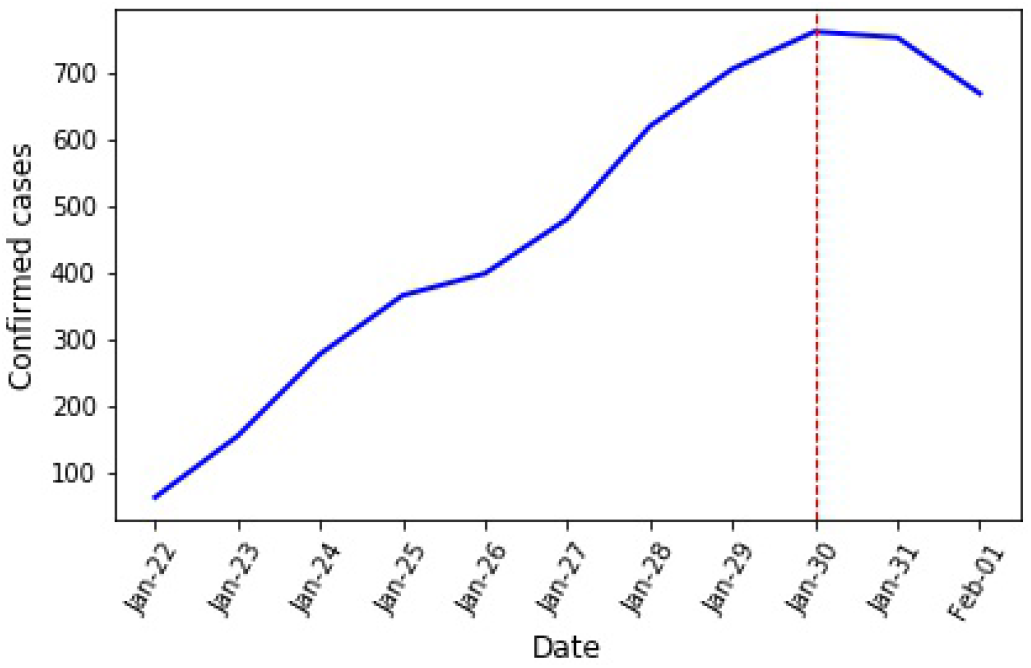
Trend of the daily confirmed cases from 01/22 to 02/01, 2020.

However, the migration raised from people returning to work after Chinese New Year on Feb 3rd undermines the continuous decline of *E*_*t*_. Since Feb 2nd, the number of daily confirmed patients in mainland China beyond Hubei Province has increased for two consecutive days, where the *E*_*t*_ on Feb 3rd has increased by 23% compared to that on Feb 2nd. It can be concluded that these fluctuations are caused by the resuming of social activities, which leads *E*_*t*_ continue to decline since Feb 4th. In many literature and media reports, Feb 3rd is used as the time point when the number of newly confirmed patients starts to decline. But considering the fact that the epidemic was already under control, here we still view Jan 30th as the first turning point.

After that, the second turning point *T*_2_, which is the time point when the number of infectious cases in hospital *N*_*t*_ starts to decline, have also been observed. Fig. 2 shows the true curves of the daily infection rate *K*_*t*_, daily removed rate *I*_*t*_, and *N*_*t*_ calculated based on the actual data from mainland China beyond Hubei from Jan 22th, 2020 to Mar 13th, 2020. It can be seen that the second turning point *T*_2_ appeared on Feb 11th, with the emergence of *K*_*t*_ < *I*_*t*_ on that day, and the number of patients in the hospital continued to decreases since then.

**Fig 2:**
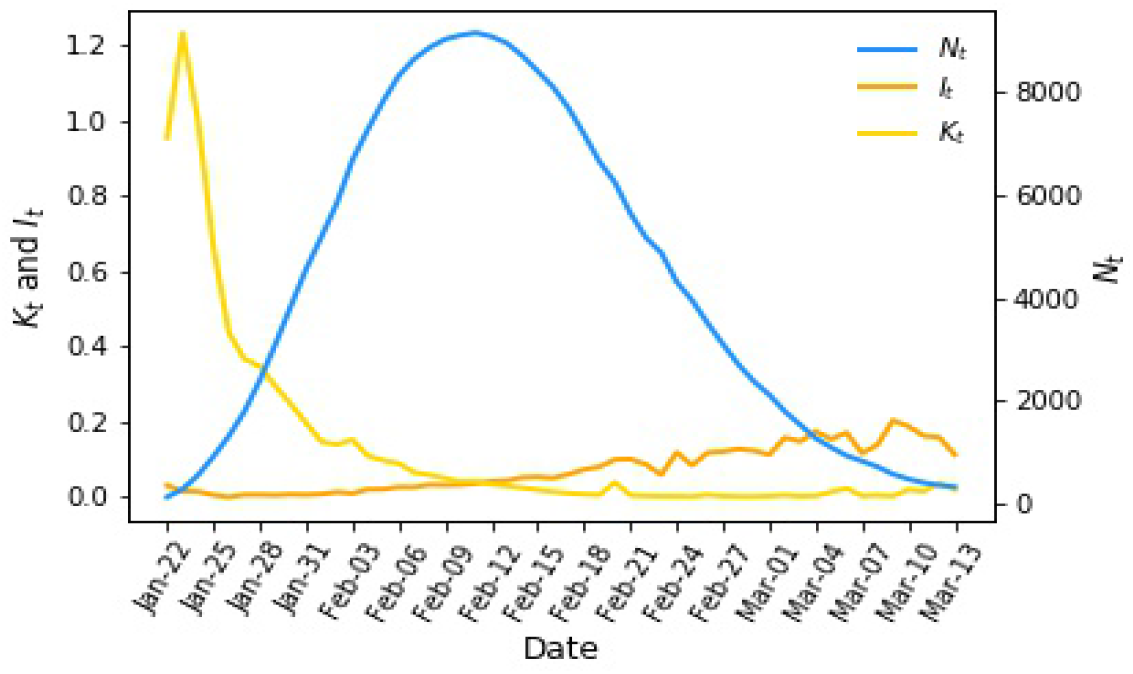
Observed *K*_*t*_, *I*_*t*_ and *N*_*t*_ of the COVID-19 from Jan 22 to Mar 13, 2020.

As for the first zero point *Z*_1_, the definition is the time when the number of daily confirmed cases is equal to zero, which is too strict for the real situation Thus, in this article, we take the criteria for cancelling travel warnings developed by the WTO during SARS as a reference, and make some adjustments to the definition of the first zero point: the time when the daily confirmed cases *E*_*t*_ continues to be less than 5 for 3 days is revised to be *Z*_1_. Then, if we exclude confirmed cases that originated from abroad, daily confirmed cases has already become less than 5 since Mar 3rd in mainland China beyond Hubei Province, thus according to our revised definition, Mar 5th is *Z*_1_. However, there were still 1,089 patients in hospital on that day. Therefore, it would still take some extra time to reach the second zero point *Z*_2_.

### 3.2 Prediction results

Starting from Jan 29th, we use the proposed forecasting method to make real-time predictions on the two turning points *T*_1_ and *T*_2_ and two “zero” points *Z*_1_ and *Z*_2_ with window size *m* = 5. The specific and predicted results are as follows.

We first conducted the proposed prediction model on Jan 29th, which indicated that the first turning point *T*_1_ would arrive on Jan 31st, i.e., *E*_*t*_ < *E*_*t*_ − 1. In reality, the first turning point did arrive on Jan 30th, which is only one day away from our predicted result.

As for the second turning point, since the true *T*_2_ occurred on Feb 11th, we summarize the frequency of the prediction results obtained with *t*_0_ varying from Jan 29th to Feb 10th, 2020 and *m* = 5 in Figure 3(a). From it we can see that the prediction of second turning point mainly concentrated in the range from Feb 9th to Feb 11th, which is consistent with the observed second turning point in reality. It is worth mentioning that we got the general information of *T*_2_ at a very early stage: we predicted on Feb 2nd that the second turning point *T*_2_ would arrive on Feb 11th, which is exactly the same as the second turning point that observed in reality. Since then, we have continuously tracked the rolling predictions, which have not yet changed much.

**Fig 3:**
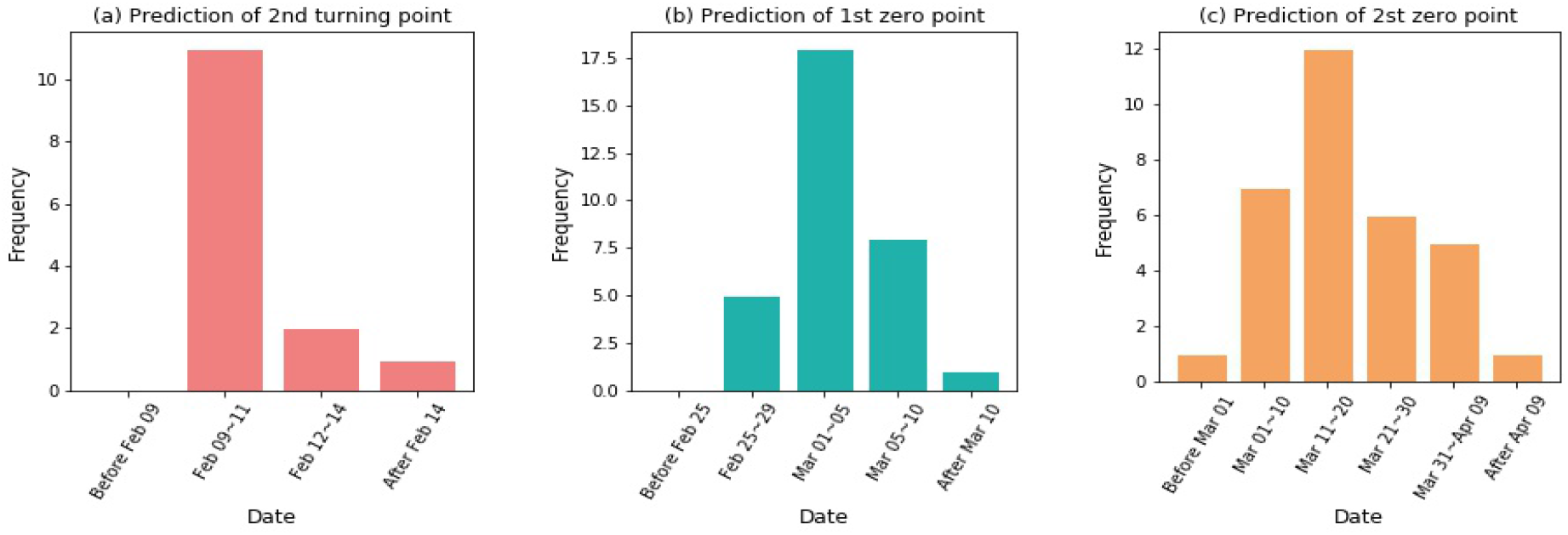
The frequency of prediction results of turning points and zero points.

Similarly, Figure 3(b) and Figure 3(c) show the frequency of the prediction results for two “zero” points obtained with *t*_0_ varying from Jan 29th to Feb 29th, 2020 and *m* = 5, respectively. Specifically, for the predicted first zero point *Z*_1_ in Figure 3(b), we divide the prediction results from these days into 5 intervals, which can be seen that the prediction results of the first zero point *Z*_1_ are mainly concentrated on Mar 1st to 5th, which is consistent with the actual result. There is also a “pessimistic” prediction as a result of the sudden fluctuation of data on Feb 3rd, which predicted that the first zero point would arrive on Mar 17th. For the predicted second zero point *Z*_2_ in Figure 3(c), it can be seen that the second zero point will be reached from mid-March to mid-April. However, there is a prediction result that *Z*_2_ will appear on May 11th, which is far away from other results. The reason for this uncommon result is that the starting point of this forecast is Jan 29th, when the epidemic situation in mainland China beyond Hubei was still in the outbreak period with *E*_*t*_ still rising, *I*_*t*_ very small, so the prediction result about the finish of the epidemic may not be accurate.

Furthermore, we also present the forecast results of the four milepost moments together with the trend of the cumulative number of infectious cases in hospital 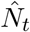 and the cumulative number of infectious 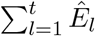 in Figure 4 when the prediction starting point *t*_0_ fixed at Jan 29th, Jan 31st, Feb 12th and Feb 26th, 2020, respectively. As can be seen from Figure 4(a), in Jan 29th, which is the very early stage of the epidemic, we predicted that the first turning point would appear on Jan 31st, which is only one day behind the actual observation. Additionally, the time of the second turning point result predicted on that day was Feb 14th, which is only 3 days away from the reality. The first zero and second zero forecast results are Mar 7th and May 11th, respectively.

**Fig 4:**
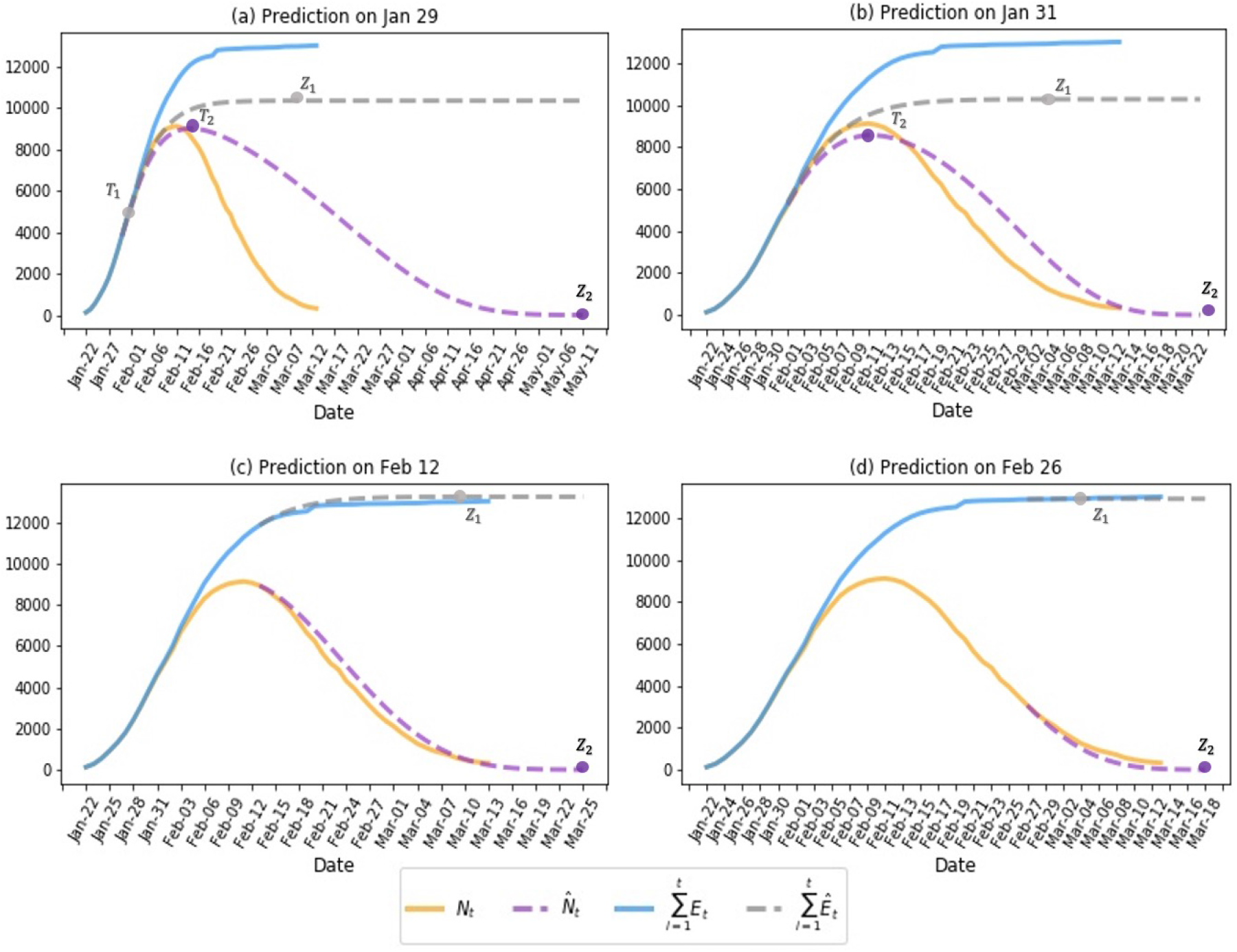
Forecasting results of the four milepost moments together with the trend of the cumulative number of infectious cases in hospital 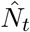 and the cumulative number of infectious 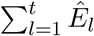 compared with their observed cases *N*_*t*_ and 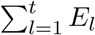 when the prediction starting point *t*_0_ fixed at Jan 29th (a), Jan 31st (b), Feb 12th (c) and Feb 26 (d), 2020, respectively.

Figure 4(b) shows the prediction results when the first turning point have already appeared, from which we can see that the prediction for *T*_2_ on Jan 31st is accurately with the second turning point possible occurring on Feb 11th. Meanwhile the first zero point and the second zero point are predicted to appear around Mar 4th and Mar 23rd, respectively.

Similarly, after the arrival of the second zero point, Figure 4(c) shows the forecast results of the first and second zero points predicted on Feb 12th, which show the forecast results for *Z*_1_ and *Z*_2_ are on Mar 9th and Mar 25th, respectively. From the fitting results, we know that our prediction of the cumulative number of patients in the hospital *N*_*t*_ and the total number of confirmed patients is very similar to the actual situation, so our prediction results are highly reliable. Finally, we also give a very recent (Feb 26th) forecast in Figure 4(d), which is similar to the results mentioned above.

### 3.3 Results with different window sizes *m*

Note that the number of *m* plays an important role in the proposed procedure, and all the results we discussed in the section 3.2 is obtained with fixed *m* = 5. In this section, we will illustrate the impact of different choice of *m* on the results, and give the empirical choice in real data analysis. Parallel to Section 3.2, here we obtain the results for the second turning point and both zero points via implementation of the proposed procedure with *m* =3, 4, and 6, respectively. And we summarize all these results for the second turning point and both zero points in Fig. 5, respectively.

**Fig 5:**
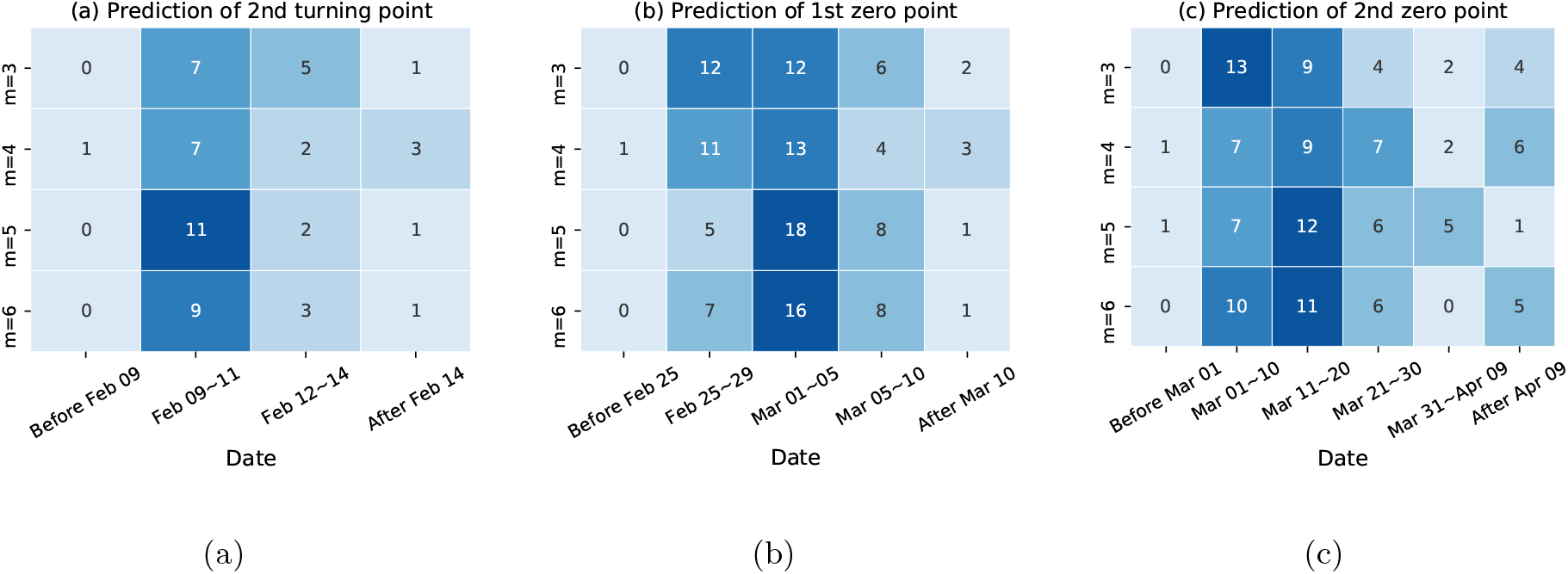
Summary of prediction for the second turning point (a), the first (b) and second (c) “zero” points with different *m*.

From Fig. 5, we can see that the most possible of forecasts for the second turning point occur around the period from Feb 9th to 11th for all choice of *m*; similar results hold for the forecast of the first zero with the most likelihood of appearance around the early March. Both results show the limited influence of *m* on the results. From Fig. 5(c), although the forecasts for the second “zero” point with different *m* seems not as good as those for the second turning point and the first zero, it vary slightly, with its occurrence from mid-March to mid-April.Overall, the choice of *m* seems not a critical value for the forecasting results, and we recommend its empirical choice from 3 to 6.

## 4 Discussion and conclusion

Focusing on the four meaningful mileposts, we put forward a simple and effective framework incorporating the effectiveness of the government control to forecast the whole process of a new unknown infectious disease in its early-outbreak. Specifically, we first propose a series of iconic indicators to characterize the extent of epidemic spread, and describe four periods of the whole process corresponding to the four meaningful milepost moments: two turning points and two “zero” points; then we develop the proposed procedure with mild and reasonable assumption, especially without relying on an assumption of epidemiological parameters for disease progression.

We examine our model with COVID-19 data in mainland China beyond Hubei province, which can detect the gross process of the epidemic at its early-outbreak. Specifically, in the first predicting task that conducted on Jan 29, the predicted date when the number of newly confirmed patients *E*_*t*_ would fall for the first time is only one day behind the observation in reality. On Feb 2nd, our model predicted that the date when the number of patients in the hospital *N*_*t*_ reaches its peak is Feb 11th, which is consistent with the real world situation. Later, the forecasting results fluctuated but were overall stable and close to the true observation. Meanwhile, we predict that the first zero point *Z*_1_ will arrive between the end of Feb and the beginning of March. And the second zero point *Z*_2_ will arrive at mid-March to mid-April. We also checked the robustness of our model under different time windows and found that the selection of the time window has little effect on the prediction of turning points. As a prediction model for the task of early warning of a new epidemic, our prediction model is proved to be quite efficient.

At present, many countries around the world are overwhelmed by the COVID-19 epidemic, which calls for global efforts. While our method is able to depict and predict the trend of an epidemic at a very early stage, it can be used to predict the current COVID-19 epidemic internationally, or any other new, unknown, explosive epidemic in the future. We believe that the prediction results of this method can provide decision support for epidemics control and intervention. It is worth noticing that, due to the short-term dependence of our method, our model may show poor performance for wildly fluctuating data. Thus, more data preprocessing methods like data smoothing need to be developed within our framework, in order to get a wider use of our method.

## Data Availability

The data used in this article is publicly available from China CDC, see details in the article.

## Acknowledgement

The authors are grateful for the financial support from the National Natural Science Foundation of China (Grant Nos. 11701023, 71843006).

## Notes

### Competing Interest Statement

The authors have declared no competing interest.

